# Neutrophil-to-Lymphocyte and Platelet-to-Lymphocyte Ratios and Cardiovascular Risk in HIV

**DOI:** 10.1101/2025.11.21.25340769

**Authors:** Jonathan N. Tobin, Yiqi Tian, Megha Khatri Arora, Takreem Ahmed, Mariam A. Siyanbola, Alondra M. Torres González, Roger Vaughan, Kevin Fiscella, Teresa H. Evering

## Abstract

**Background:** People living with HIV (PLWH) are at increased risk for cardiovascular disease (CVD); current prediction models underestimate risk for PLWH. We assessed whether adding neutrophil-to-lymphocyte ratio (NLR) and platelet-to-lymphocyte ratio (PLR), inflammatory biomarkers derived from routine complete blood counts to the Atherosclerotic Cardiovascular Disease (ASCVD) risk score improves prediction of hard ASCVD events in PLWH.

**Methods:** Retrospective cohort study (2009–2019) using electronic health records from the Bronx Regional Health Information Organization. Adults aged 40–79 were included: 7,556 PLWH and 22,545 demographically matched people without HIV (PLWoH) (1:3); those with pre-existing CVD were excluded. Incident hard ASCVD events were identified using ICD-9/10 codes. Cox proportional hazards models assessed associations between HIV status, NLR and PLR quartiles, ASCVD risk score, and CVD events.

**Results:** The final cohort had a mean age of 56 years; 45% female, 65% Black/African American, 39% Hispanic/Latino. PLWH had higher rates of hard ASCVD events than PLWoH (6.2% vs 4.7%, p<0.001). In the fully adjusted Cox model, HIV-positive status was independently associated with 36% higher risk (HR 1.358; 95% CI 1.218–1.515). The highest NLR quartile was associated with increased risk (HR 1.518; 95% CI 1.400–1.646); higher PLR quartiles showed an inverse association. Adding NLR and PLR modestly improved discrimination in the overall cohort (concordance 0.661 to 0.680) and PLWoH (0.670 to 0.691), but not in PLWH (0.640 to 0.638).

**Conclusions:** NLR and PLR provide modest incremental predictive value in the overall cohort and PLWoH but not in PLWH. HIV-specific risk stratification tools remain needed.

**CLINICAL PERSPECTIVE:** *What Is New?:*

- In a large, diverse cohort of 30,101 adults from the Bronx, New York, including 7,556 people living with HIV, the addition of neutrophil-to-lymphocyte ratio (NLR) and platelet-to-lymphocyte (PLR) ratio to the ASCVD risk score produced a modest improvement in cardiovascular risk discrimination in the overall cohort but did not meaningfully improve discrimination among people living with HIV specifically.

*What Are the Clinical Implications?:*

- Ratio-derived routine, low-cost inflammatory biomarkers derived from the complete blood count may offer incremental value for cardiovascular risk stratification in general populations but appear insufficient to capture the excess cardiovascular risk conferred by HIV infection.
- These findings reinforce the need for HIV-specific cardiovascular risk assessment tools and continued emphasis on aggressive management of modifiable risk factors in people living with HIV, a population in which established risk calculators are known to underestimate cardiovascular risk.

## INTRODUCTION

Age- and sex-adjusted deaths from cardiovascular disease (CVD) among people living with HIV (PLWH) are significantly higher than in the general population and extend to those in younger age groups ^1–5^. This excess risk reflects multiple factors, including greater burden of traditional CVD risk factors in PLWH ^6,7^. However, chronic inflammation, marked by increased proinflammatory cytokines, immune cell activation, and elevated coagulation markers ^8–10^, plays a key role in the pathogenesis of coronary artery disease (CAD) ^11^. Persisting during effective combination antiretroviral therapy, residual immune activation is also believed to play an important role in the premature development of CVD in PLWH ^1,3,12,13^.

Current CVD prediction models underestimate CVD risk in PLWH ^13,14^. Inflammatory markers such as high-sensitivity C-reactive protein (hs-CRP) have proven strongly predictive of CVD events and mortality in cohort studies in the general population ^15–19^. However, hs-CRP is not routinely measured. The neutrophil-to-lymphocyte ratio (NLR) and platelet-to-lymphocyte ratio (PLR) represent low-cost, routinely available putative biomarkers with potential utility to improve CVD risk prediction ^20,21^. These markers of systemic inflammation are easily calculated from routine blood count parameters.

The NLR is a valuable prognostic marker in various populations ^22,23^. In the general population, the NLR has demonstrated utility in stratifying all-cause and CVD mortality ^24,25^. This prognostic value extends to specific groups, as reported in a systematic review and meta-analysis in which the NLR predicted all-cause mortality and cardiovascular events in patients with chronic kidney disease ^26^. A retrospective cohort study of 1,454 patients with CVD and type 2 diabetes mellitus showed that over a seven-year period, individuals with a lower NLR (≤2.5) showed significantly better survival compared to those with a higher NLR (>2.5) ^27^. Other inflammatory markers have also shown similar potential. A cross-sectional study (n=1146) found elevated PLR significantly associated with metabolic-syndrome and higher CRP levels ^28^.

Recently, more complex inflammatory indices have emerged ^29^. The systemic immune-inflammation index (SII), comprised of platelet count x neutrophil/lymphocyte ratio, was demonstrated to predict the occurrence and prognosis of cardiovascular events in patients with CAD who had undergone percutaneous coronary intervention (PCI) ^30^. A 2022 meta-analysis and systematic review found that higher SII was significantly associated with an increased CVD risk across almost all CVD subtypes, though the quality of evidence was assessed as generally low ^31^. A prospective cohort study of 13,026 obese adults from the National Health and Nutrition Examination Survey (NHANES) found that both the systemic inflammatory response index (SIRI), calculated as (neutrophil × monocyte)/lymphocyte, and SII were independent risk factors for all-cause and CVD mortality ^32^.

Retrospective studies have also explored how the NLR and PLR perform in combination. In 300 individuals with acute myocardial infarction, overall survival (OS) and progression-free survival (PFS) decreased with increasing PLR and NLR values ^33^. Similarly, in patients hospitalized with acute heart failure (HF) (n=321), NLR and mean platelet volume-to-lymphocyte ratio (MPVLR) at admission independently predicted adverse cardiovascular events, HF rehospitalization, in-hospital mortality, and composite outcomes ^34^.

The NLR and PLR have also shown utility in assessing CVD risk and overall mortality in PLWH ^35^. In an Italian retrospective cohort study of PLWH, NLR was an independent predictor of CVD risk (HR 3.05 for NLR≥ 1.2) ^36^ and both markers have been independently associated with all-cause mortality in PLWH ^37^. Additionally, in a case-control study comparing sex- and age-matched PLWH with and without hypertension, while NLR was among the inflammatory markers found to be significantly higher in those in the hypertension group, PLR differences were not statistically significant ^38^.

An important subset of CVD in PLWH is cerebrovascular disease. While the lifespan of PLWH has increased considerably in the era of effective combination antiretroviral therapy (cART), HIV-associated cerebrovascular disease remains highly prevalent ^39,40^. As this population ages, there is increasing overlap between HIV-associated cerebrovascular dysfunction and aging-related co-morbidities, such as hypertension and diabetes ^41,42^. HIV-1 enters the central nervous system (CNS) early in infection ^43^ and may influence the subsequent response of intra- and extra-cranial arteries to inflammation and other vascular risk factors ^44^. Residual systemic inflammation despite cART may influence the severity of HIV-associated vasculopathy and ischemic stroke severity ^45,46^. In addition, the relative contribution of neuroinflammation and atherosclerosis to cerebral vessel wall remodeling is unclear ^47^. Whether NLR and PLR improve CVD risk prediction in large, diverse cohorts of PLWH in the United States remains unclear.

Given evidence of shared mechanisms of disease pathogenesis between cardiac and cerebrovascular disease, we hypothesized that the NLR and PLR would positively correlate with hard ASCVD event incidence (defined to include both cardiac and cerebrovascular outcomes) in PLWH, and that addition of these markers to baseline ASCVD risk scores would significantly improve CVD risk prediction in PLWH.

## METHODS

### Data Availability

The authors will make available all the data required to replicate the analysis described in this manuscript upon reasonable request and execution of a Data Use Agreement. Readers interested in the data obtained from the Bronx RHIO can email Megha Khatri Arora, MBA for data requests.

### Study Population Selection

We identified all adults (age ≥ 18 years) from the Bronx Regional Health Information Organization (“Bronx RHIO”), a large health information exchange (HIE) containing complete electronic health records (EHRs) covering most of healthcare provided to the 1.4 million Bronx NY residents. From the Bronx RHIO database (n=2,751,664), we identified 34,140 adults with an HIV-1 diagnosis (PLWH) and 2,717,524 without an HIV diagnosis (PLWoH) between January 1, 2009 and December 31, 2019. Data extracted included demographics (age, sex, race/ethnicity, insurance status), diagnostic (HIV status, hypertension, diabetes, smoking), laboratory (blood pressure, cholesterol, complete blood count (CBC)), and prescription data.

We applied sequential exclusion criteria to both groups to arrive at our final analysis cohort (Figure 1 – STROBE Diagram). After excluding those missing race, gender, blood pressure readings, cholesterol values, and CBC panel results, we had 15,174 PLWH and 360,917 PLWoH. For each eligible PLWH, three demographically matched controls without HIV were randomly selected using a validated algorithm developed by Rockefeller University Biostatistics based on demographics and location of care ^48^. To minimize bias, the HIV diagnosis date of each PLWH served as the index date for the corresponding matched PLWoH controls. Following 1:3 HIV-positive to HIV-negative matching and removing laboratory outliers (see Supplementary Methods), 11,434 PLWH and 31,882 PLWoH remained. We further restricted the cohort to participants aged 40-79 years consistent with the eligibility range of the Pooled Cohort Equations (excluding 8,455 participants) and excluding participants whose HIV diagnosis date occurred after the index date (n=1,815) to ensure appropriate temporal ordering of exposure and outcome. After excluding those with pre-existing CVD at or before the index date (n=2,945), our final analysis cohort included 7,556 PLWH and 22,545 demographically matched PLWoH (total n=30,101). This exclusion criterion ensures that all predictor measurements preceded incident CVD outcomes for every participant in the analytic cohort.

**Figure 1.**
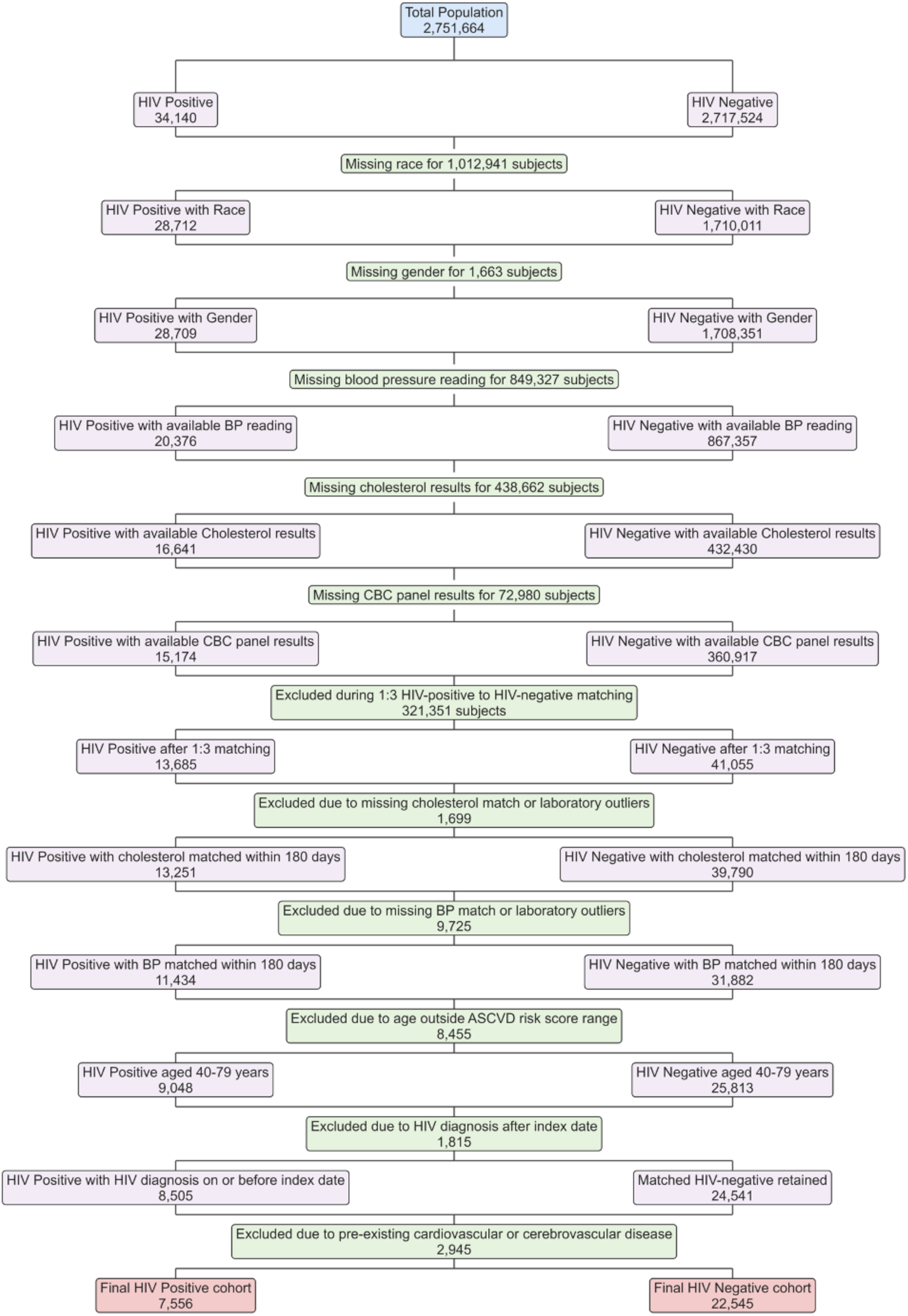
STROBE Flow Diagram of Study Population Selection. Flow diagram showing the selection of the study population from the initial database to the final analysis cohort. The process details sequential exclusions for missing data, laboratory outliers, age outside the ASCVD risk score eligibility range, HIV diagnosis after the index date, and pre-existing cardiovascular disease.

### Calculation of 10-year ASCVD Risk Scores, NLR and PLR

ASCVD risk scores were calculated using the American Heart Association (AHA) ASCVD Risk Estimator Plus, incorporating laboratory tests, vital signs, diagnoses, and medications ^49^. We filtered lab records for ‘Total cholesterol’ and ‘HDL cholesterol,’ removing implausible outliers. Given the varying time points and less frequent occurrence of cholesterol measurements compared to blood pressure records, we used deterministic matching by patient ID and date, with the cholesterol test date serving as the anchor date for risk score calculation. Additional laboratory variables were aligned to the index date using a ±180-day matching window, selecting the temporally closest available measurement for each patient. Participants were not excluded if additional laboratory values were unavailable within this window; missing laboratory values were addressed using multiple imputation ^50,51^. Antihypertensive treatment, diabetes, and smoking status were determined from medication lists and ICD 9/10 diagnostic codes.

Baseline NLR and PLR were calculated from the baseline CBC with differential. NLR was calculated by dividing the absolute neutrophil count (ANC) by the absolute lymphocyte count (ALC). PLR was calculated by dividing the absolute platelet count (APC) by the ALC. We excluded extreme values likely reflecting data entry errors or biologically implausible results (neutrophil values ≤0.5 or ≥30 × 10⁹/L, platelet counts ≤10 or ≥1000 × 10⁹/L, and lymphocyte counts ≤0.5 or ≥15 × 10⁹/L). These thresholds were based on clinical laboratory reference ranges and established data quality standards ^52,53^. NLR and PLR values were categorized into quartiles (Table S2).

### Additional Variables

HIV viral load data excluded lab test names related to ‘phenotype,’ ‘antibody,’ ‘Geno,’ ‘multisport,’ ‘interpretation,’ ‘screen,’ and ‘western blot’. Log-transformed values were converted to copies/ml using base 10 exponentiation. HIV viral load was categorized into two groups: ‘<200 copies/ml’ (“virologically suppressed”) and ‘≥200 copies/ml’ (“not virologically suppressed”).

### Outcomes

#### Primary Outcome

The primary outcome was incident hard ASCVD events over an 11-year follow-up period (2009– 2019), identified using prespecified ICD-9/10 codes, defined as myocardial infarction or stroke (see Table S1) ^54–58^.

#### Secondary Outcomes

Secondary outcomes included comparisons of incident CVD diagnoses between PLWH and PLWoH, stratified by NLR and PLR, with adjustment for baseline ASCVD risk. We also evaluated the predictive value of NLR and PLR for incident CVD events by assessing whether their inclusion improved risk prediction beyond baseline ASCVD risk scores.

### Statistical Analysis

Baseline demographic and clinical characteristics were summarized and stratified by HIV status. Continuous variables were reported as means and standard deviations, and categorical variables as counts and percentages. Comparisons used Wilcoxon rank-sum tests for continuous variables and Pearson’s Chi-squared or Fisher’s exact tests for categorical variables.

To assess the relationship between HIV status, inflammatory markers and cardiovascular outcomes, we constructed four Cox proportional hazards regression models with progressive complexity: Model 1 (ASCVD risk score and HIV status), Model 2 (Model 1 + NLR quartile), Model 3 (Model 1 + PLR quartile), and Model 4 (Model 1 + both NLR and PLR quartiles). Follow-up time was defined from each participant’s index date to the earliest of incident hard ASCVD event or administrative censoring on December 31, 2019. Hazard ratios (HRs) with 95% confidence intervals (CIs) were reported. Cox models were fit separately in the overall cohort, among PLWH, and among PLWoH. Model performance was evaluated using concordance statistics and likelihood ratio tests (LRTs) ^59^ to assess improvements in fit. Parallel logistic regression analyses were conducted as a secondary analysis, with results presented in Table S3. Analyses were conducted in R version 4.4.2, with statistical significance defined as p < 0.05.

As this was a retrospective cohort study using all eligible participants from the Bronx RHIO, the sample size was determined by data availability. With 30,101 participants and 1,519 incident hard ASCVD events (event rate 5.0%), the study had >80% power to detect hazard ratios of approximately 1.15–1.20 at a two-sided α of 0.05.

### Missing Data

Missing values for complete blood count components (lymphocyte, neutrophil, and platelet counts) were handled using multiple imputation by chained equations with predictive mean matching, generating 20 imputed datasets prior to calculation of NLR and PLR and subsequent statistical analyses ^51,60^.

### Regulatory Considerations

The study was approved by a central institutional review board (IRB) (BRANY), with reliance agreements in place across the collaborating institutions (Weill Cornell Medicine, BronxRHIO, and Clinical Directors Network). Informed consent requirements were waived per IRB determination for this retrospective database study using de-identified data.

## RESULTS

### Study Population Characteristics

The analysis included 30,101 participants (7,556 PLWH and 22,545 PLWoH) meeting all eligibility criteria (STROBE, Figure 1). Table 1 presents baseline characteristics, which were generally well-matched on demographics (age, sex, race, ethnicity, and borough of residence) by design. Social deprivation index (SDI) scores and insurance coverage are also summarized in Table 1. The overall mean age was 55.6 ± 8.2 years, with PLWH and PLWoH being similar in age (55.4 ± 8.2 vs 55.7 ± 8.3 years, p<0.005). The cohort was predominantly male (55%) and Black or African American (65%), with substantial Hispanic/Latino representation (39%). The majority of participants resided in the Bronx (94%), reflecting the Bronx RHIO source population.

**Table 1.** Baseline Characteristics of Study Participants by HIV Status.

| Variable | Overall N = 30,101<sup>1</sup> | HIV+ N = 7,556<sup>1</sup> | HIV- N = 22,545<sup>1</sup> | p-value<sup>2</sup> |
| --- | --- | --- | --- | --- |
| Age (years) | 55.63 (8.23) | 55.39 (8.19) | 55.70 (8.25) | 0.005 |
| Gender |  |  |  | 0.7 |
| Female | 13,596 (45%) | 3,396 (45%) | 10,200 (45%) |  |
| Male | 16,505 (55%) | 4,160 (55%) | 12,345 (55%) |  |
| Race |  |  |  | 0.5 |
| American Indian Or Alaska Native | 107 (0.4%) | 22 (0.3%) | 85 (0.4%) |  |
| Asian | 110 (0.4%) | 28 (0.4%) | 82 (0.4%) |  |
| Black Or African American | 19,593 (65%) | 4,870 (64%) | 14,723 (65%) |  |
| Native Hawaiian Or Other Pacific Islander | 229 (0.8%) | 61 (0.8%) | 168 (0.7%) |  |
| Other Race | 3,413 (11%) | 853 (11%) | 2,560 (11%) |  |
| White | 6,649 (22%) | 1,722 (23%) | 4,927 (22%) |  |
| Ethnicity |  |  |  | 0.5 |
| Hispanic | 11,684 (39%) | 2,967 (39%) | 8,717 (39%) |  |
| Not Hispanic | 18,286 (61%) | 4,560 (60%) | 13,726 (61%) |  |
| Unknown | 131 (0.4%) | 29 (0.4%) | 102 (0.5%) |  |
| Borough |  |  |  | 0.3 |
| Bronx | 28,194 (94%) | 7,059 (93%) | 21,135 (94%) |  |
| Other NYC | 1,907 (6.3%) | 497 (6.6%) | 1,410 (6.3%) |  |
| SDI Score | 97.27 (7.55) | 97.23 (7.58) | 97.28 (7.53) | 0.3 |
| Insurance |  |  |  | <0.001 |
| Government Aid | 28,634 (95%) | 7,271 (96%) | 21,363 (95%) |  |
| Other/Unknown | 1,335 (4.4%) | 267 (3.5%) | 1,068 (4.7%) |  |
| Private Insurance | 122 (0.4%) | 18 (0.2%) | 104 (0.5%) |  |
| Uninsured | 10 (<0.1%) | 0 (0%) | 10 (<0.1%) |  |
| Lymphocyte Count | 2.08 (0.84) | 2.01 (0.87) | 2.11 (0.82) | <0.001 |
| Neutrophil Count | 4.16 (2.50) | 3.78 (2.43) | 4.28 (2.51) | <0.001 |
| Platelet Count | 241.33 (77.11) | 224.30 (78.96) | 247.03 (75.63) | <0.001 |
| NLR | 2.39 (2.25) | 2.28 (2.21) | 2.42 (2.26) | <0.001 |
| PLR | 132.03 (68.04) | 129.95 (73.94) | 132.73 (65.93) | <0.001 |
| Total Cholesterol | 182.95 (42.63) | 175.63 (43.00) | 185.40 (42.22) | <0.001 |
| HDL Cholesterol | 51.78 (15.57) | 49.81 (15.86) | 52.44 (15.41) | <0.001 |
| LDL Cholesterol | 104.99 (36.87) | 97.59 (35.91) | 107.47 (36.86) | <0.001 |
| Systolic BP | 131.54 (20.68) | 127.62 (20.08) | 132.85 (20.71) | <0.001 |
| Diastolic BP | 79.30 (29.90) | 76.92 (20.20) | 80.09 (32.48) | <0.001 |
| Antihypertensive Treatment | 8,930 (30%) | 2,487 (33%) | 6,443 (29%) | <0.001 |
| Diabetes | 12,460 (41%) | 2,948 (39%) | 9,512 (42%) | <0.001 |
| Smoker | 4,872 (16%) | 1,857 (25%) | 3,015 (13%) | <0.001 |
| ASCVD Risk (%) | 10.63 (10.39) | 10.33 (10.26) | 10.73 (10.43) | <0.001 |
| Cardiovascular Disease | 642 (2.1%) | 215 (2.8%) | 427 (1.9%) | <0.001 |
| Cerebrovascular Disease | 991 (3.3%) | 295 (3.9%) | 696 (3.1%) | <0.001 |
| Cardiovascular or Cerebrovascular Disease | 1,519 (5.0%) | 468 (6.2%) | 1,051 (4.7%) | <0.001 |
| HIV viral load (copies/mL) |  |  |  |  |
| <200 copies/ml |  | 1,316 (52%) | NA |  |
| >=200 copies/ml |  | 1,218 (48%) |  |  |
| CD4 count category (cells/μL) |  |  |  |  |
| <200 |  | 1,168 (19%) | NA |  |
| 200-349 |  | 978 (16%) |  |  |
| 350-499 |  | 996 (17%) |  |  |
| >=500 |  | 2,853 (48%) |  |  |

<sup>1</sup>Mean (SD); n (%)
<sup>2</sup>Wilcoxon rank sum test; Pearson's Chi-squared test; Fisher's exact test
<sup>3</sup>Percentages for viral load and CD4 count are calculated among PLWH with available measurements within 180 days of the index date (n = 2,534 and 5,995, respectively)

### Clinical and Laboratory Characteristics

Smoking was substantially more prevalent among PLWH compared to PLWoH (25% vs 13%), while diabetes was slightly less common in PLWH (39% vs 42%). Blood pressure parameters were lower in PLWH, with mean systolic blood pressure of 127.6 vs 132.9 mmHg and diastolic blood pressure of 76.9 vs 80.1 mmHg, though a higher proportion of PLWH were on antihypertensive treatment (33% vs 29%). Lipid profiles also differed, with PLWH having lower mean total cholesterol (175.6 vs 185.4 mg/dL), LDL cholesterol (97.6 vs 107.5 mg/dL), and HDL cholesterol (49.8 vs 52.4 mg/dL) levels compared to PLWoH. All between-group differences were statistically significant (p<0.001). Mean ASCVD risk scores were similar in PLWH (10.3% vs 10.7%).

Among PLWH, 52% had HIV viral load <200 copies/mL. 19% had CD4 T cell counts <200 cells/µL, indicating significant immunosuppression.

### Immune Inflammatory Markers

We examined inflammatory markers beyond traditional ASCVD risk factors for additional risk stratification. The inflammatory markers showed distinct patterns between groups: The absolute neutrophil and lymphocyte counts were lower in PLWH compared to PLWoH (3.78 vs 4.28 ×10³/µL and 2.01 vs 2.11 ×10³/µL, respectively; p<0.001 for both). PLWH had lower mean NLR values compared to PLWoH (2.28 vs 2.42, p<0.001). PLR also differed significantly between groups, with PLWH having slightly lower values than PLWoH (129.95 vs 132.73, p<0.001).

### Primary Outcome

Incident hard ASCVD events were observed in 1,519 (5.0%) participants, with higher prevalence among PLWH (468; 6.2%) than PLWoH (1,051; 4.7%, p<0.001), suggesting increased cardiovascular risk. Within these hard ASCVD events, cerebrovascular disease specifically occurred in 295 PLWH (3.9%) compared to 696 PLWoH (3.1%, p<0.001). Median follow-up time was 23.4 months (IQR 11.1-34.9) among PLWH and 23.2 months (IQR 11.7-34.7) among PLWoH (p=0.025), a difference not likely to be clinically meaningful.

### Multi-variable Modeling

Sequential modeling revealed independent associations between HIV status and hard ASCVD events across all model specifications (Tables 2a and 2b). In Model 1, HIV-positive status was associated with 34% higher risk of hard ASCVD events after adjusting for ASCVD risk score (HR 1.334; 95% CI: 1.196–1.487). ASCVD risk score was also significantly associated with the primary outcome (HR per 1% increase: 1.039; 95% CI: 1.034–1.044). Adding NLR quartiles (Model 2) led to a substantial improvement in model fit (likelihood ratio χ² 527.4 vs 427.1), with the highest NLR quartile strongly associated with increased risk (HR 1.388; 95% CI: 1.292–1.491). Adding PLR quartiles (Model 3) did not meaningfully improve model fit (likelihood ratio χ² 427.2 vs 427.1), and PLR quartiles were not independently associated with hard ASCVD events in this model (HR 1.007; 95% CI: 0.942–1.077).

**Table 2a.** Cox Proportional Hazards Regression Models Predicting Incident Hard ASCVD Events in the Overall Cohort: Main Effects.

| Variable | Model 1 | Model 2 | Model 3 | Model 4 |
| --- | --- | --- | --- | --- |
| ASCVD Risk Score | 1.039 (1.034–1.044) | 1.035 (1.030–1.040) | 1.039 (1.034–1.044) | 1.034 (1.029–1.039) |
| HIV+ Status | 1.334 (1.196–1.487) | 1.366 (1.225–1.524) | 1.334 (1.196–1.488) | 1.358 (1.218–1.515) |
| NLR quartile | - | 1.388 (1.292–1.491) | - | 1.518 (1.400–1.646) |
| PLR quartile | - | - | 1.007 (0.942–1.077) | 0.835 (0.775–0.900) |

**Table 2b.** Cox Proportional Hazards Regression Models Predicting Incident Hard ASCVD Events in the Overall Cohort: Model Performance.

| Metric | Model 1 | Model 2 | Model 3 | Model 4 |
| --- | --- | --- | --- | --- |
| Concordance | 0.661 | 0.677 | 0.661 | 0.680 |
| Likelihood Ratio $\chi^2$ | 427.1 (df=3) | 527.4 (df=5) | 427.2 (df=5) | 554.1 (df=7) |

The fully adjusted model including both NLR and PLR quartiles (Model 4) achieved the best model fit (likelihood ratio χ² 554.1). Patients in the highest NLR quartile had 52% higher risk of hard ASCVD events (HR: 1.518; 95% CI: 1.400–1.646), while those in the highest PLR quartile had 17% lower risk (HR: 0.835; 95% CI: 0.775–0.900), compared to the lowest quartile. Model concordance improved from 0.661 in Model 1 to 0.680 in Model 4.

When analyses were stratified by HIV status, NLR remained independently associated with hard ASCVD events among PLWH in the fully adjusted model (HR 1.375; 95% CI: 1.180–1.603), with a similar pattern observed among PLWoH (HR 1.567; 95% CI: 1.424–1.725). PLR showed an inverse association in both groups. However, model concordance did not improve among PLWH with the addition of NLR and PLR quartiles (0.640 in Model 1 vs 0.638 in Model 4), while modest improvement was observed among PLWoH (0.670 in Model 1 to 0.691 in Model 4) (Tables 3a and 3b; PLWoH results in Table S4).

**Table 3a.** Cox Proportional Hazards Regression Models Predicting Incident Hard ASCVD Events Among People Living with HIV: Main Effects.

| Variable | Model 1 | Model 2 | Model 3 | Model 4 |
| --- | --- | --- | --- | --- |
| ASCVD Risk Score | 1.034 (1.023–1.045) | 1.032 (1.021–1.043) | 1.034 (1.023–1.045) | 1.030 (1.019–1.042) |
| NLR quartile | - | 1.237 (1.078–1.418) | - | 1.375 (1.180–1.603) |
| PLR quartile | - | - | 0.923 (0.807–1.054) | 0.800 (0.690–0.928) |

**Table 3b.** Cox Proportional Hazards Regression Models Predicting Incident Hard ASCVD Events Among People Living with HIV: Model Performance.

| **Metric** | **Model 1** | **Model 2** | **Model 3** | **Model 4** |
| --- | --- | --- | --- | --- |
| Concordance | 0.640 | 0.636 | 0.636 | 0.638 |
| Likelihood Ratio $\chi^2$ | 81.35 (df=2) | 91.78 (df=4) | 83.12 (df=4) | 102.0 (df=6) |

## DISCUSSION

In this combined cohort of PLWH and demographically matched PLWoH, NLR and PLR quartiles were independently associated with incident hard ASCVD events after adjustment for baseline ASCVD risk score. Adding NLR and PLR to the ASCVD risk score resulted in modest improvement in model discrimination in the overall cohort, with concordance increasing from 0.661 to 0.680 in Cox models. The highest NLR quartile was strongly associated with increased cardiovascular risk in the fully adjusted model (HR 1.518, 95% CI 1.400-1.646), while higher PLR quartiles showed an inverse association with risk, a finding contrary to our original hypothesis, that is discussed further below. When stratified by HIV status, the addition of NLR and PLR did not meaningfully improve discrimination among PLWH specifically, with concordance remaining essentially unchanged from 0.640 to 0.638, while greater improvement was observed among PLWoH, with concordance increasing from 0.670 to 0.691. While these findings suggest that NLR and PLR provide limited incremental value for CVD risk prediction in PLWH beyond established risk scores, the need for improved risk stratification in this high-risk population remains important. Data from the HIV Outpatient Study ^61^, where 8.5% of participants experienced incident CVD events over an 11-year period, underscore this unmet clinical need.

Despite being similar in age, with more favorable traditional risk factors (lower blood pressure, cholesterol levels, and slightly lower diabetes prevalence) than PLWoH, PLWH showed significantly higher hard ASCVD event rates. The notable exception was smoking prevalence, markedly higher in PLWH (25% vs 13%), consistent with previous studies showing 2-3 times higher smoking rates in this population ^62^. The 36% higher risk of events in our fully adjusted model indicates substantial residual risk beyond what traditional ASCVD risk scores capture, supporting previous findings that current risk calculators underestimate risk in PLWH.

Our findings align with Quiros-Roldan et al., who reported HR 3.05 for NLR≥1.2 as a CVD predictor in PLWH ^36^. Similarly, Raffetti et al. found both NLR and PLR independently associated with all-cause mortality in PLWH ^37^. We observed that PLWH had lower NLR and lower PLR values compared to matched controls. This pattern likely reflects hematologic effects of HIV infection and combination antiretroviral therapy, which suppress bone marrow function and reduce absolute neutrophil counts disproportionately to lymphocyte counts, yielding a lower calculated NLR despite an underlying pro-inflammatory state ^63–67^. Wang et al. reported similar findings with a lower Systemic Immune-Inflammation Index in PLWH, hypothesizing hematologic alterations from cART ^68^. Importantly, a lower absolute NLR in this population does not indicate reduced inflammation, as persistent systemic immune activation has been documented in PLWH despite viral suppression ^69^. HIV- and cART-induced hematologic changes may also moderate the predictive performance of NLR and PLR in PLWH, consistent with our observation that addition of these markers improved model discrimination in PLWoH but not in PLWH. More recently, a case-control study found NLR to be significantly higher in hypertensive compared to normotensive PLWH in univariate analysis, while PLR differences were not statistically significant ^38^.

The inverse association between PLR and hard ASCVD events observed in our cohort was contrary to our original hypothesis, and we cannot fully explain the direction of this association with the available data. However, two features of this cohort may contextualize the divergence from prior literature. First, an inverse association between PLR and hypertension has been previously reported, with a 2024 Zambian study finding higher PLR inversely associated with hypertension in PLWH (adjusted OR 0.98, 95% CI 0.97-0.99, p=0.01), suggesting that the inverse PLR finding in our cohort is not an isolated observation ^70^. Second, in this cohort both platelet and lymphocyte counts were lower in PLWH than PLWoH (platelets 224.3 vs 247.0 K/µL, lymphocytes 2.01 vs 2.11 K/µL, both p<0.001), consistent with HIV-associated hematologic suppression affecting both cell lines. The biological meaning of PLR in this population may therefore differ from that in prior studies reporting a positive PLR-CVD association. The mechanistic basis for the inverse PLR-CVD association warrants investigation in larger prospective studies of similar populations.

The mechanistic basis for persistent CVD risk in PLWH involves chronic inflammation that persists despite effective antiretroviral therapy, with HIV-infected macrophages in atherosclerotic plaques secreting inflammatory cytokines that promote endothelial dysfunction and immune cell recruitment ^10^. HIV entry into the CNS ^43^ may impact vascular responses to inflammation ^44^. Residual systemic inflammation may influence the severity of HIV-associated vasculopathy and ischemic stroke ^45,46^ though the relative contribution of neuroinflammation and atherosclerosis to cerebral vessel wall remodeling is unclear ^47^.

Study strengths include a large, diverse, geographic population-based cohort with extended follow-up, real-world EHR data from underserved urban populations, and robust sample size enabling findings broadly applicable to similar populations. However, several limitations exist. As an observational study, we cannot establish causation between inflammatory markers and outcomes. Missing data on key covariates resulted in exclusion of large numbers of individuals, potentially introducing selection bias and limiting generalizability. NLR and PLR levels were measured at a single timepoint, and potential changes in these biomarkers over time were not evaluated. HIV viral load data were available for only a subset of PLWH (n=2,534, 34%), limiting the power of viral load sensitivity analyses. Additionally, viral load was measured at a single timepoint, and whether sustained virological suppression modifies the NLR-CVD and PLR-CVD associations over time could not be assessed. Death was not ascertained in this cohort due to the nature of the EHR data source; participants without a recorded hard ASCVD event were censored at December 31, 2019 regardless of vital status, which may have led to underestimation of event rates. The date of HIV acquisition is unknown for all participants, as the recorded diagnosis date reflects clinical identification rather than true date of infection incidence, precluding a precise adjustment for duration of HIV infection. Although the study adjusted for numerous potential confounders, residual confounding due to unmeasured variables cannot be entirely excluded. Finally, the mechanistic pathways linking these inflammatory markers to cardiovascular outcomes remain unclear, though causal pathway analysis by Angkananard et al. demonstrated that NLR’s cardiovascular impact is both direct and mediated through traditional risk factors ^71^.

These findings have important clinical implications. Recent REPRIEVE trial results demonstrated that Pooled Cohort Equations significantly underpredicted CVD events in women and Black or African American men with HIV in high-income countries, highlighting limitations of current risk prediction tools and need for HIV-specific assessment approaches ^72^. Although NLR and PLR were independently associated with hard ASCVD events and provided modest incremental discriminative value in the overall cohort and among PLWoH, they did not meaningfully improve CVD risk discrimination among PLWH specifically. This may reflect reduced statistical power in the smaller PLWH subgroup or may indicate that the excess CVD risk in PLWH is driven by HIV-specific mechanisms beyond systemic inflammation captured by NLR and PLR, such as immune activation, viral replication, and cART effects, that these markers do not fully capture, and/or HIV-related modification of the relationship between these inflammatory markers and CVD incidence. The clinical utility of these markers for risk stratification in PLWH therefore remains uncertain and warrants further investigation in larger prospective studies. The persistently elevated CVD risk observed in PLWH in this and other studies underscores the importance of aggressive screening and preventive approaches such as the “Million Hearts” initiative or ABCS (**A**spirin, **B**lood pressure, **C**holesterol, **S**moking cessation) strategies in this high-risk population ^73,74^, with continued efforts needed to develop and validate HIV-specific risk stratification tools.

The PREVENT equations, recently developed by the American Heart Association, were not incorporated in our study, as our analyses were pre-specified using the ASCVD Risk Estimator Plus prior to PREVENT’s publication. Recent evidence suggests that PREVENT may exacerbate underprediction of cardiovascular risk in PLWH compared to the Pooled Cohort Equations ^75,76^, reinforcing the continued relevance of the ASCVD Risk Estimator Plus as the comparator model in studies of this population.

Future research should examine these associations in other, larger populations, particularly prospective studies with sufficient power to evaluate NLR and PLR as risk stratification tools specifically in PLWH. Studies examining time-varying inflammatory markers and viral load, and their interaction effects over the course of HIV treatment, would help clarify whether these associations hold in contemporary treatment settings where virological suppression rates are substantially higher. Most importantly, establishing their role in guiding therapeutic interventions for CVD prevention in PLWH remains a critical research priority.

## Acknowledgements

The authors thank Ken Rapkin of the Campbell Foundation and Kathryn Miller of BronxRHIO for their support of this work.

## Supplementary Materials

Supplementary Methods

Table S1. ICD-9 and ICD-10 Diagnostic Codes Used to Define Hard ASCVD Events

Table S2. Quartile Cutoff Values for NLR and PLR

Table S3. Stratified Logistic Regression Models Predicting Incident Hard ASCVD Events: Main Effects and Model Performance by HIV Status

Table S4. Cox Proportional Hazards Regression Models Predicting Incident Hard ASCVD Events Among People Without HIV: Main Effects and Model Performance

Table S5. Sensitivity Analyses: Association Between Inflammatory Markers and Incident Hard ASCVD Events by Viral Load Status Among People Living with HIV

Table S6. Interaction Term Results for Viral Load Status and Inflammatory Markers Among People Living with HIV

