## Supplementary Data for "Neutrophil-to-Lymphocyte and Platelet-to-Lymphocyte Ratios and Cardiovascular Risk in HIV"

**Supplementary Online Content**

**Supplementary Methods**


Study Population Selection Process

Laboratory Data Processing

Data Management & Data Analysis


**Supplementary Tables**


Table S1: ICD-9 and ICD-10 Diagnostic Codes Used to Define Hard ASCVD Events

Table S2: Quartile Cutoff Values for NLR and PLR

Table S3: Stratified Logistic Regression Models Predicting Incident Hard ASCVD Events: Main Effects and Model Performance by HIV Status

Table S4: Cox Proportional Hazards Regression Models Predicting Incident Hard ASCVD Events Among People Without HIV: Main Effects and Model Performance

Table S5: Sensitivity Analyses: Association Between Inflammatory Markers and Incident Hard ASCVD Events by Viral Load Status Among People Living with HIV

Table S6: Interaction Term Results for Viral Load Status and Inflammatory Markers Among People Living with HIV

**Supplementary Methods**

Study Population Selection Process

Electronic health record data were obtained from the Bronx Regional Health Information Organization (RHIO), including laboratory measurements, blood pressure records, diagnoses, medications, demographic characteristics, and insurance information. All measurements were restricted to January 1, 2009 through December 31, 2019. Participants with available demographic information were first identified (n=54,740; HIV-positive=13,685; HIV-negative=41,055). Individuals with available cholesterol measurements matched within ±180 days were retained after exclusion of those with missing cholesterol matches or laboratory outliers (n=53,041; HIV-positive=13,251; HIV-negative=39,790). This was followed by retention of individuals with blood pressure records matched within ±180 days after exclusion of those with missing blood pressure matches or laboratory outliers (n=43,316; HIV-positive=11,434; HIV-negative=31,882). The cohort was restricted to individuals aged 40–79 years at baseline (n=34,861; HIV-positive=9,048; HIV-negative=25,813), consistent with the eligibility range of the pooled cohort ASCVD risk equations. To ensure appropriate temporal ordering, participants whose HIV diagnosis date occurred after the index date were excluded (n=33,046; HIV-positive=8,505; HIV-negative=24,541). To construct an inception cohort, individuals with a hard ASCVD diagnosis prior to the index date were excluded, resulting in a final analytic cohort of 30,101 participants (HIV-positive=7,556; HIV-negative=22,545).

Baseline clinical measurements were anchored on an index date defined through a stepwise matching procedure using lipid and blood pressure records. Total cholesterol and HDL cholesterol measurements were first evaluated; if both measurements were available on the same day, the earliest such date was selected; otherwise, the earliest total cholesterol measurement with an HDL cholesterol value within ±180 days was identified. Blood pressure measurements were subsequently aligned to these lipid records. If total cholesterol and systolic blood pressure were available on the same day, the earliest such date was retained; otherwise, the nearest systolic blood pressure measurement within ±180 days of the cholesterol date was selected. The resulting laboratory date was defined as the index date for each participant. Additional laboratory variables, including LDL cholesterol, diastolic blood pressure, lymphocyte count, neutrophil count, platelet count, and HIV viral load (for PLWH), were aligned to the index date using the same ±180-day matching window. When multiple measurements were available on the same date, values were averaged. Participants were not excluded if these additional laboratory values were unavailable within the matching window; missing laboratory values were addressed using multiple imputation.

Inflammatory biomarkers were derived from complete blood count data. The neutrophil-to-lymphocyte ratio (NLR) was calculated as neutrophil count divided by lymphocyte count, and the platelet-to-lymphocyte ratio (PLR) was calculated as platelet count divided by lymphocyte count. The 10-year ASCVD risk score was calculated using the pooled cohort equations incorporating age, sex, race, total cholesterol, HDL cholesterol, systolic blood pressure, antihypertensive treatment, diabetes, and smoking status. Antihypertensive treatment was defined as initiation of antihypertensive medication on or before the index date based on medication name and National Drug Code records. Diabetes and smoking status were defined using diagnosis records prior to baseline. HIV viral load values were cleaned, converted from log values when necessary, and categorized as <200 copies/mL or ≥200 copies/mL. The primary outcome was incident hard ASCVD events defined as myocardial infarction or stroke identified using prespecified ICD-9 and ICD-10 diagnosis codes. For each participant, the earliest qualifying event date was recorded. Missing values for complete blood count components (lymphocyte, neutrophil, and platelet counts) were handled using multiple imputation by chained equations with predictive mean matching, generating 20 imputed datasets prior to calculation of NLR and PLR and subsequent statistical analyses.

Laboratory Data Processing

We filtered neutrophils, lymphocytes, and platelets from approximately 2 gigabytes of laboratory data and standardized the units to 'K/µL'. Since EHRs are based on manual data entry and may contain typos or erroneous values, such as 0 due to rounding issues, we excluded extreme values that were likely to reflect data entry errors or biologically implausible results.

Data Management & Data Analysis

Using the PCORnet Common Data Model (CDM), BronxRHIO programmers transformed EHR clinical, laboratory, medication, procedures and utilization data into a standardized set of variable definitions with a common data dictionary. Data were queried for International Classification of Diseases, Clinical Modification Ninth and Tenth Revision (ICD-9-CM and ICD-10-CM) diagnostic codes from January 1, 2009 through December 31, 2019. All data from Health Information Exchange (HIE) participants are stored in a document-based NoSQL system that uses elastic search engine technology to query records. Records were retrieved for all diagnostic codes in any care setting. Python programming language was then used to extract and cleanse pertinent data from medical records. Data on selected elements, including hospital name, medical record number, date of diagnosis, visit type, and date of birth, were extracted in csv format and exported for analysis.

**Supplementary Tables**

**Table S1. ICD-9 and ICD-10 Diagnostic Codes Used to Define Hard ASCVD Events**

| **Outcome** | **Included ICD-9 codes** | **Included ICD-10 codes** | **Exclusions** | **Supporting references** |
|---|---|---|---|---|
| Acute Myocardial Infarction | 410.x0, 410.x1 | I21.x, I22.x | Exclude 410.x2 (subsequent episode), old MI (412, I25.2), angina (413.x, I20.x), chronic ischemic heart disease (414.x, I25.x) | Goff et al, Circulation, 2014; McCormick et al, PLoS One, 2014; Patel et al, CMAJ Open, 2015 |
| Stroke | 430, 431, 433.x1, 434.x1, 436 | I63.x | Exclude stenosis without infarction (433.x0, 434.x0), TIA (435.x, G45.x), stroke sequelae (I69.x), chronic cerebrovascular disease (437.x, I67.x) | Goff et al, Circulation, 2014; Kokotailo et al, Stroke, 2005; Thigpen et al, Circ Cardiovasc Qual Outcomes, 2015 |
| Excluded non-hard ASCVD conditions | 412, 413.x, 414.x, 435.x, 437.x, 440.x, 443.x | I20.x, I25.x, G45.x, I67.x, I69.x, I70.x, I73.9, I74.x | Angina, chronic CAD, TIA, PAD, stroke sequelae, abnormal ECG, ECMO history | Goff et al, Circulation, 2014 |

**Table S2. Quartile Cutoff Values for NLR and PLR**

| Percentile | NLR | PLR |
|---|---|---|
| 0% | 0.099 | 5.000 |
| 25% | 1.211 | 88.974 |
| 50% | 1.778 | 117.500 |
| 75% | 2.700 | 156.698 |
| 100% | 39.917 | 1485.000 |

**Table S3. Stratified Logistic Regression Models Predicting Incident Hard ASCVD Events: Main Effects and Model Performance by HIV Status**

**Overall Cohort: Main Effects (Logistic Regression)**

| Variable | Model 1 | Model 2 | Model 3 | Model 4 |
|---|---|---|---|---|
| ASCVD Risk | 1.040 (1.036–1.044) | 1.037 (1.034–1.041) | 1.040 (1.036–1.044) | 1.036 (1.032–1.040) |
| HIV+ Status | 1.383 (1.234–1.548) | 1.418 (1.265–1.588) | 1.368 (1.220–1.532) | 1.395 (1.244–1.562) |
| NLR quartile Q2 | - | 1.109 (0.938–1.312) | - | 1.188 (1.004–1.408) |
| NLR quartile Q3 | - | 1.311 (1.116–1.542) | - | 1.502 (1.271–1.778) |
| NLR quartile Q4 | - | 1.935 (1.667–2.252) | - | 2.405 (2.032–2.850) |
| PLR quartile Q2 | - | - | 0.784 (0.676–0.910) | 0.703 (0.604–0.818) |
| PLR quartile Q3 | - | - | 0.853 (0.737–0.987) | 0.680 (0.583–0.792) |
| PLR quartile Q4 | - | - | 0.959 (0.833–1.104) | 0.619 (0.527–0.727) |

**Overall Cohort: Model Performance**

| Metric | Model 1 | Model 2 | Model 3 | Model 4 |
|---|---|---|---|---|
| AUC | 0.659 | 0.673 | 0.661 | 0.679 |
| Cutoff | 0.044 | 0.047 | 0.047 | 0.047 |
| Sensitivity | 0.647 | 0.644 | 0.602 | 0.659 |
| Specificity | 0.586 | 0.619 | 0.640 | 0.612 |
| Balanced Accuracy | 0.617 | 0.631 | 0.621 | 0.635 |

**People Living with HIV: Main Effects (Logistic Regression)**

| Variable | Model 1 | Model 2 | Model 3 | Model 4 |
|---|---|---|---|---|
| ASCVD Risk | 1.033 (1.025–1.040) | 1.032 (1.024–1.039) | 1.033 (1.025–1.040) | 1.031 (1.023–1.038) |
| NLR quartile Q2 | - | 1.032 (0.782–1.361) | - | 1.101 (0.832–1.456) |
| NLR quartile Q3 | - | 1.181 (0.899–1.549) | - | 1.340 (1.010–1.777) |
| NLR quartile Q4 | - | 1.451 (1.122–1.879) | - | 1.778 (1.332–2.376) |
| PLR quartile Q2 | - | - | 0.884 (0.684–1.141) | 0.834 (0.642–1.079) |
| PLR quartile Q3 | - | - | 0.803 (0.613–1.047) | 0.701 (0.529–0.923) |
| PLR quartile Q4 | - | - | 0.867 (0.673–1.115) | 0.656 (0.493–0.872) |

**People Living with HIV: Model Performance (Logistic Regression)**

| Metric | Model 1 | Model 2 | Model 3 | Model 4 |
|---|---|---|---|---|
| AUC | 0.620 | 0.625 | 0.624 | 0.630 |
| Cutoff | 0.055 | 0.056 | 0.059 | 0.055 |
| Sensitivity | 0.641 | 0.658 | 0.558 | 0.658 |
| Specificity | 0.557 | 0.550 | 0.644 | 0.535 |
| Balanced Accuracy | 0.599 | 0.604 | 0.601 | 0.597 |

**People Without HIV: Main Effects (Logistic Regression)**

| Variable | Model 1 | Model 2 | Model 3 | Model 4 |
|---|---|---|---|---|
| ASCVD Risk | 1.043 (1.038–1.048) | 1.040 (1.035–1.044) | 1.043 (1.038–1.047) | 1.038 (1.034–1.043) |
| NLR quartile Q2 | - | 1.178 (0.953–1.457) | - | 1.266 (1.023–1.569) |
| NLR quartile Q3 | - | 1.415 (1.157–1.737) | - | 1.635 (1.326–2.021) |
| NLR quartile Q4 | - | 2.227 (1.848–2.697) | - | 2.802 (2.271–3.469) |
| PLR quartile Q2 | - | - | 0.744 (0.620–0.892) | 0.646 (0.536–0.777) |
| PLR quartile Q3 | - | - | 0.874 (0.734–1.041) | 0.662 (0.550–0.796) |
| PLR quartile Q4 | - | - | 0.997 (0.841–1.182) | 0.594 (0.489–0.723) |

**People Without HIV: Model Performance (Logistic Regression)**

| Metric | Model 1 | Model 2 | Model 3 | Model 4 |
|---|---|---|---|---|
| AUC | 0.669 | 0.684 | 0.670 | 0.692 |
| Cutoff | 0.043 | 0.037 | 0.044 | 0.047 |
| Sensitivity | 0.588 | 0.744 | 0.589 | 0.604 |
| Specificity | 0.664 | 0.540 | 0.678 | 0.679 |
| Balanced Accuracy | 0.626 | 0.642 | 0.633 | 0.641 |

Odds ratios (95% confidence intervals) are presented for each variable across four sequential models, stratified by HIV status. Model 1 includes ASCVD risk score and HIV status in the overall cohort, or ASCVD risk score only in HIV-stratified models. Model 2 adds NLR quartile. Model 3 adds PLR quartile. Model 4 includes both NLR and PLR quartiles. NLR and PLR are entered as categorical quartile variables with the lowest quartile as the reference group. Hard ASCVD events were defined as incident myocardial infarction or stroke identified using prespecified ICD-9 and ICD-10 diagnostic codes.

**Table S4. Cox Proportional Hazards Regression Models Predicting Incident Hard ASCVD Events Among People Without HIV: Main Effects and Model Performance**

**Main Effects**

| Variable | Model 1 | Model 2 | Model 3 | Model 4 |
|---|---|---|---|---|
| ASCVD Risk | 1.040 (1.034–1.046) | 1.036 (1.030–1.042) | 1.040 (1.034–1.046) | 1.035 (1.029–1.041) |
| NLR quartile | - | 1.437 (1.320–1.564) | - | 1.567 (1.424–1.725) |
| PLR quartile | - | - | 1.030 (0.953–1.114) | 0.842 (0.772–0.919) |

**Model Performance**

| Metric | Model 1 | Model 2 | Model 3 | Model 4 |
|---|---|---|---|---|
| Concordance | 0.670 | 0.688 | 0.670 | 0.691 |
| Likelihood Ratio $\chi^2$ | 329.8 (df=2) | 424.1 (df=4) | 330.8 (df=4) | 441.9 (df=6) |

Hazard ratios (95% confidence intervals) are presented for each variable across four sequential models among PLWoH. Model 1 includes ASCVD risk score only. Model 2 adds NLR quartile. Model 3 adds PLR quartile. Model 4 includes both NLR and PLR quartiles. NLR and PLR are entered as categorical quartile variables with the lowest quartile as the reference group. Hard ASCVD events were defined as incident myocardial infarction or stroke identified using prespecified ICD-9 and ICD-10 diagnostic codes. Follow-up time was defined from each participant's index date to the earliest of incident hard ASCVD event or censoring on December 31, 2019. Concordance is equivalent to the C-statistic for survival models and reflects overall model discrimination. Likelihood ratio chi-square tests the overall significance of each model relative to a null model. df, degrees of freedom.

**Table S5. Sensitivity Analyses: Association Between Inflammatory Markers and Incident Hard ASCVD Events by Viral Load Status Among People Living with HIV**

| **(A) Multivariable Logistic Regression Among PLWH with HIV Viral Load** | | |
|---|---|---|
| **Variable** | **OR** | **95% CI** |
| Intercept | 0.058 | (0.038 - 0.085) |
| ASCVD risk | 1.038 | (1.025 - 1.051) |
| VL status: Suppressed | 0.751 | (0.544 - 1.034) |
| NLR quartile: Q2 vs Q1 | 1.037 | (0.643 - 1.664) |
| NLR quartile: Q3 vs Q1 | 1.441 | (0.896 - 2.315) |
| NLR quartile: Q4 vs Q1 | 1.559 | (0.953 - 2.553) |
| PLR quartile: Q2 vs Q1 | 0.595 | (0.367 - 0.940) |
| PLR quartile: Q3 vs Q1 | 0.588 | (0.363 - 0.935) |
| PLR quartile: Q4 vs Q1 | 0.701 | (0.442 - 1.105) |

| **(B) Multivariable Cox Proportional Hazards Model Among PLWH with HIV Viral Load** | | |
|---|---|---|
| Variable | HR | 95% CI |
| ASCVD risk | 1.027 | (1.007 - 1.046) |
| VL status: Suppressed | 0.785 | (0.577 - 1.067) |
| NLR quartile: Q2 vs Q1 | 1.031 | (0.654 - 1.626) |
| NLR quartile: Q3 vs Q1 | 1.425 | (0.906 - 2.243) |
| NLR quartile: Q4 vs Q1 | 1.520 | (0.949 - 2.434) |
| PLR quartile: Q2 vs Q1 | 0.639 | (0.407 - 1.003) |
| PLR quartile: Q3 vs Q1 | 0.628 | (0.400 - 0.988) |
| PLR quartile: Q4 vs Q1 | 0.765 | (0.495 - 1.182) |

| (C) Stratified Logistic Regression by Viral Load Status Among PLWH | | | | |
|---|---|---|---|---|
| | < 200 copies/ml (Suppressed) | | >=200 copies/ml (Unsuppressed) | |
| **Variable** | **OR** | **95% CI** | **OR** | **95% CI** |
| Intercept | 0.050 | (0.028 - 0.085) | 0.048 | (0.028 - 0.080) |
| ASCVD risk | 1.027 | (1.008 - 1.046) | 1.048 | (1.028 - 1.067) |
| NLR quartile: Q2 vs Q1 | 1.529 | (0.781 - 3.017) | 0.704 | (0.347 - 1.384) |
| NLR quartile: Q3 vs Q1 | 1.667 | (0.806 - 3.448) | 1.395 | (0.739 - 2.624) |
| NLR quartile: Q4 vs Q1 | 1.742 | (0.825 - 3.703) | 1.447 | (0.748 - 2.796) |
| PLR quartile: Q2 vs Q1 | 0.200 | (0.074 - 0.457) | 1.180 | (0.647 - 2.127) |
| PLR quartile: Q3 vs Q1 | 0.580 | (0.303 - 1.078) | 0.601 | (0.283 - 1.207) |
| PLR quartile: Q4 vs Q1 | 0.662 | (0.334 - 1.287) | 0.766 | (0.408 - 1.437) |

| (D) Stratified Cox Proportional Hazards Models by Viral Load Status Among PLWH | | | | |
|---|---|---|---|---|
| | < 200 copies/ml (Suppressed) | | >=200 copies/ml (Unsuppressed) | |
| **Variable** | **HR** | **95% CI** | **HR** | **95% CI** |
| ASCVD risk | 1.014 | (0.986 - 1.043) | 1.035 | (1.009 - 1.062) |
| NLR quartile: Q2 vs Q1 | 1.544 | (0.809 - 2.946) | 0.700 | (0.364 - 1.347) |
| NLR quartile: Q3 vs Q1 | 1.725 | (0.861 - 3.454) | 1.314 | (0.722 - 2.388) |
| NLR quartile: Q4 vs Q1 | 1.729 | (0.843 - 3.547) | 1.434 | (0.767 - 2.681) |
| PLR quartile: Q2 vs Q1 | 0.214 | (0.089 - 0.515) | 1.254 | (0.719 - 2.188) |
| PLR quartile: Q3 vs Q1 | 0.611 | (0.333 - 1.122) | 0.652 | (0.328 - 1.297) |
| PLR quartile: Q4 vs Q1 | 0.716 | (0.377 - 1.360) | 0.821 | (0.453 - 1.488) |

Results are presented for four complementary analyses examining the influence of viral load status on the association between NLR, PLR, and incident hard ASCVD events among PLWH. Panel A presents multivariable logistic regression with viral load status included as a covariate. Panel B presents Cox proportional hazards regression with viral load status included as a covariate. Panel C presents stratified logistic regression by virological suppression status. Panel D presents stratified Cox proportional hazards regression by virological suppression status. Viral load suppression was defined as VL <200 copies/mL. Analyses were restricted to PLWH with available viral load data (n=2,534). NLR and PLR are entered as categorical quartile variables with the lowest quartile as the reference group.

**Table S6. Interaction Term Results for Viral Load Status and Inflammatory Markers**

**Cox Model - NLR Interaction**

| Term | HR | 95% CI | P value |
|---|---|---|---|
| VL>=200 × NLR Q2 | 0.539 | (0.215, 1.347) | 0.186 |
| VL>=200 × NLR Q3 | 0.740 | (0.307, 1.783) | 0.502 |
| VL>=200 × NLR Q4 | 0.654 | (0.283, 1.510) | 0.320 |

**Cox Model - PLR Interaction**

| Term | HR | 95% CI | P value |
|---|---|---|---|
| VL>=200 × PLR Q2 | 0.808 | (0.284, 2.297) | 0.689 |
| VL>=200 × PLR Q3 | 1.002 | (0.404, 2.484) | 0.996 |
| VL>=200 × PLR Q4 | 0.684 | (0.315, 1.487) | 0.338 |

**Logistic Model - NLR Interaction**

| Term | OR | 95% CI | P value |
|---|---|---|---|
| VL>=200 × NLR Q2 | 0.514 | (0.198, 1.306) | 0.164 |
| VL>=200 × NLR Q3 | 0.989 | (0.399, 2.450) | 0.981 |
| VL>=200 × NLR Q4 | 0.851 | (0.356, 2.029) | 0.716 |

**Logistic Model - PLR Interaction**

| Term | OR | 95% CI | P value |
|---|---|---|---|
| VL>=200 × PLR Q2 | 5.220 | (1.895, 16.179) | 0.002 |
| VL>=200 × PLR Q3 | 0.939 | (0.368, 2.346) | 0.894 |
| VL>=200 × PLR Q4 | 1.111 | (0.500, 2.496) | 0.797 |

Results of formal interaction testing between viral load status (VL ≥200 vs <200 copies/mL) and NLR and PLR quartiles in both logistic and Cox proportional hazards models among PLWH with available viral load data (n=2,534). Interaction terms are presented for each NLR and PLR quartile separately. Statistically significant interaction terms indicate that the association between the inflammatory marker and incident hard ASCVD events differs meaningfully by virological suppression status.
